# Satisfaction with perioperative anesthesia care among general surgery patients at a secondary level hospital, Dodoma-Tanzania

**DOI:** 10.1101/2025.08.01.25332672

**Authors:** Janeth S. Masuma, Alphoncina Kagaigai, Amani Albert Ulimali, Ibenzi Njile, Peter Mbele, Frank Sandi, Amani Anaeli

**Affiliations:** School of Public Health and Social Sciences, Muhimbili University of Health and Allied Sciences, P.O Box 65001, Dar es Salaam, Tanzania, institutional website: www.muhas.ac.tz; Department of Development Studies, School of Public Health and Social Sciences, Muhimbili University of Health and Allied Sciences, P.O. Box 65454, Dar es Salaam Tanzania; Department of Anesthesiology, Muhimbili Orthopedic Institute (MOI), P.O. Box 65474 Dar-es-salaam, Tanzania; Department of Surgery, Dodoma Regional Referral Hospital (DRRH), P.O. Box 904, Dodoma, Tanzania; Department of Ophthalmology and visual sciences, School of medicine and Dentistry, The University of Dodoma, P.O. Box 395, Dodoma, Tanzania

**Keywords:** Satisfaction, perioperative anesthesia care, Dodoma-Tanzania

## Abstract

**Introduction:** This study aims to determine patients’ satisfaction with perioperative anesthesia care and its associated factors among patients planned for surgery at Dodoma Regional Referral Hospital (DRRH), Tanzania.

**Methods:** An analytical Cross-sectional study of 425 perioperative patients admitted at DRRH using an interview form modified from the Leiden Perioperative Care Patient Satisfaction questionnaire, was conducted. Data was collected using an Open data Kit (ODK) tool. Perioperative factors were analysed per specific objective and where applicable percentage, mean and standard deviations were used. Satisfaction was measured using demarcation threshold formula. Association between variables was tested via Chi-square test for categorical variables. The chi-square, odds ratio, 95% confidence interval, and p-value were computed to identify associated factors and determine the strength of the association. P-value of 0.05 was used for statistical significance. Independent factors in univariate analysis with a P-value < 0.2 were included in the multivariate analysis. SPSS computer program version 20 was utilized for analysis.

**Results:** About 65% (64.9%) of respondents were satisfied with the perioperative anaesthesia care services, Overall mean (SD) satisfaction score being 69 (8.0). Postoperative surgical pain, time spent in the recovery room and staff consideration of patient’s privacy in theatre were predictive factors for patient satisfaction after perioperative anesthesia service with AOR (95% CI) P value of 0.2 (1.36-3.08) 0.003, 4.9(2.4-9.9) <0.001, and 3.2 (1.1–9.46) 0.029 respectively.

**Discussion and Conclusion:** Overall patient satisfaction with perioperative anesthesia services was moderately low. Key factors influencing satisfaction were preoperative visits, postoperative pain, ASA status, patient privacy, and recovery room time. Future research should focus on patients’ willingness to return for surgery and explore the underlying reasons behind satisfaction to improve perioperative care quality.

## INTRODUCTION

Patient satisfaction plays a critical role in assessing the effectiveness and quality of healthcare services, especially in perioperative anesthesia care, which is key to continuous hospital care improvement [1]. It reflects the degree to which patient needs and expectations are met, and it relies on subjective assessments [1,2]. Patient satisfaction influences various aspects of behavior, including healthcare resource use, treatment adherence, and relationships with healthcare providers [3,4]. Providing patient-centered care that addresses patients’ well-being physically, mentally, and spiritually contributes to a positive healthcare environment [1,5].

Key factors that shape patient satisfaction include interpersonal relationships, healthcare provider skills, and the accessibility and convenience of services, all of which are influenced by institutional policies.[6] The Donabedian model underscores the importance of assessing structure, process, and outcomes to improve healthcare quality[7]. Satisfaction metrics, such as those in perioperative anesthesia care, are crucial as they reflect patients’ subjective experiences and perceptions, driving improvements in healthcare delivery systems. Globally, patient satisfaction rates vary, with developed countries reporting rates of 90-95%, while in developing nations, the range spans from 95% to below 50%, impacting healthcare-seeking behaviors and potentially contributing to increased morbidity and mortality[8,9]

The perioperative phase involves complex care coordination among various professionals, making patient satisfaction vital not only for surgical outcomes but for the entire care continuum. Satisfaction is shaped by factors like preoperative education, pain management, and communication. In contemporary surgeries, various anesthetic types are frequently used, including general anesthesia, regional anesthesia, and possibly a combination of both [1–3]. In order to determine risk factors related to anesthesia and surgery, as well as intraoperative management and postoperative adverse impact therapy, anesthesiologists are extremely crucial. Conducting a thorough assessment of patient satisfaction with perioperative-anesthesia care is a crucial element for quality control and ongoing enhancement of hospital services[10].This study aims to assess perioperative anesthesia care satisfaction among general surgery patients at Dodoma Regional Referral Hospital in Tanzania, providing evidence to support improved care practices globally.

## Material and methods

### Study Design

An analytical Cross-sectional study design, which is hospital based, was conducted at Dodoma Regional Referral Hospital (DRRH) in Dodoma from May to June 2024. All eligible participants from the study population were interviewed to enquire about their satisfaction with the perioperative anaesthesia care services.

### Study Area

The study was done at Dodoma Referral Regional Hospital, which is the closest hospital in the region with 3,084,000 million people. Patients were recruited from general surgical wards where patients who need surgical care are admitted. Recruitment was done from 1st May to June 2024. When the decision for surgery is made, attending clinician/surgeon admits the patient to the general surgical wards, for review including preanesthetic workout including choices for anaesthesia to ensure his/her fitness. While still in the ward for preoperative workout, nurses/clinicians do administer written consent after explanations on the details of the surgery to be conducted. All cleared fit for surgery patients were then taken to operating rooms according to the list.

Patients received in the operating room, get reassessed by anesthetist and necessary premedication given depending on the assessment. Also, patients’ concerns are addressed during the operation, whether through communication with the patients (If given regional anaesthesia) and/or physiological examination (If under GA). Anesthetic care continues even after operation, and till patients are discharged, so as to address any anticipated concerns i.e pain, post-operative nausea and vomiting etc. During the post-operative period is when patients meeting criteria were enrolled in the study after consenting.

### Study population

Post-operative patients admitted at Dodoma Regional Referral Hospital and at least 24 hours have passed and have not exceeded 48 hours, since their general surgical procedure.

### Study variables

Outcome (dependent) variables was patient satisfaction category (i.e Satisfied or dissatisfied). Independent variables consisted of social demographics, preoperative, intraoperative and postoperative variables.

### Sample size and sampling technique

A sample size of 420 was required for this study. A simple random sampling technique was used in this study, where patients in the ward were assigned numbers, and a lottery method was applied to randomly selected participants, ensuring equal chances for every individual to be included. Seven participants from general surgical wards were interviewed daily for 60 days, allowing for broad coverage and minimal bias. Informed consent was obtained after explaining the research objectives, voluntary participation, and expected time commitment, and an interview survey was used to assess perioperative satisfaction with anesthesia care.

### Data collection methods

For this study, data were collected using an interview form based on the Leiden Perioperative Care Patient Satisfaction Questionnaire (LPPSq), a standardized tool for assessing perioperative patient satisfaction with anesthesia care. The LPPSq has been validated in similar settings, including Ethiopia and Eritrea, making it suitable for use in Tanzania[11,12].

The questionnaire covered three sections: patient demographics, preoperative data, and modified LPPSq factors, with responses rated according to LPPSq guidelines for pre-, intra- and postoperative experiences, as well as willingness to return. Data collection was conducted face-to-face with patients in the post-surgical wards at Dodoma Regional Referral Hospital, and additional medical information, such as ASA status and premedication, was obtained from medical records.

### Data management and analysis

Data was collected through a mobile-based application. Subsequently, they were transferred to Microsoft Excel for organization and cleansing. Routine checks for data accuracy and resolution of any discrepancies or issues were conducted.

Data analysis was done using the Statistical Package for Social Sciences (SPSS) version 20. It was analysed per each objective. For the demographic characteristics, the categorical variables (i.e gender, education, marital status) were described using proportions and continuous variables (i.e Age) were described in terms of means or media.

The overall patients’ satisfaction score was determined based on the Demarcation threshold formula using three LPPSQ domains (information provision −6 questions, fear and concern - 6 questions and staff patient realationship-14 questions) which incorporates for all stages of preoperative, intraoperative and post-operative.

Demarcation threshold formula= (Total highest score – total lowest score)/2 + total lowest score. For preoperative, intraoperative and postoperative factors were analyzed in terms of frequencies and proportions, against the outcome (i.e satisfied and dissatisfied) considering a dichotomized 5-point Likert scale, where satisfied were those considered to have scored greater than or equal to the cut point based on the demarcation threshold formula[13].

The willingness to return to the same hospital following the perioperative anaesthesia care was determined in frequencies and proportions. Categorical variables were compared using Chi-Square, while Continuous variables were compared using Z-Test, P values < 0.05 was considered significant. Dependent outcome variables were graded as satisfied and dissatisfied. Significant independent factors in univariate analysis were included in the multivariate analysis. STROBE checklist for observational studies was used to write this manuscript[14].

## RESULTS

### Demographic characteristics

A total of 425 perioperative general surgical patients admitted to Dodoma Regional Referral Hospital were interviewed using the adopted Leiden Perioperative Care Patient Satisfaction (LPPSq) interview guide. The respondence rate was 100%. The majority (32.7%) were in the age range between 25 to 34 years. The mean (SD) age was 34 (16.7) years. Among these, 53.2% were males. In terms of educational attainment, 45.2% had a secondary education level. The majority (77.2%) resided in urban areas. 58.8% of the participants were married. Table 1 below illustrates this information.

**Table 1:** Demographic characteristics of the study population(n=425)

| Demographic variables | Frequency | Percentage (%) |
| --- | --- | --- |
| **Sex** |  |  |
| Male | 226 | 53.2 |
| Female | 199 | 46.8 |
| **Age group (in years)** |  |  |
| <25 | 101 | 23.8 |
| 25-34 | 139 | 32.7 |
| 35-44 | 70 | 16.5 |
| 45-54 | 50 | 11.7 |
| >54 | 65 | 15.3 |
| **Level of education** |  |  |
| No formal education | 4 | 0.9 |
| Primary | 114 | 26.8 |
| Secondary | 192 | 45.2 |
| College/University | 115 | 27.1 |
| **Occupation** |  |  |
| Employed | 107 | 25.2 |
| Self employed | 165 | 38.8 |
| Unemployed | 98 | 23.1 |
| Other | 55 | 12.9 |
| **Residence/Setting** |  |  |
| Urban | 328 | 77.2 |
| Rural | 97 | 22.8 |
| **Marital status** |  |  |
| Married | 250 | 58.8 |
| Single | 160 | 37.6 |
| Divorced | 10 | 2.4 |
| Separated | 2 | 0.5 |
| Widowed | 3 | 0.7 |
| Others |  |  |

### Overall patient satisfaction with perioperative anesthesia care among surgical patients admitted at Dodoma Regional Referral Hospital, Tanzania

In this study, three components of satisfaction were used to calculate the overall mean satisfaction of patients with perioperative anaesthesia care, including information provision, Fear and concern and patient-staff relationship. The overall mean (SD) satisfaction score was 69 (8.0).

Using the threshold score from the demarcation formula, the majority 64.94% were satisfied and 35.06% were dissatisfied, figure1 below illustrates the information. Patients who scored ≥70 were categorized as satisfied by using the demarcation threshold formula.

When preoperative factors were distributed across patients’ satisfaction with perioperative anesthesia care, the results show that among the 425 study respondents, the majority (70.1%) of those who had a preoperative anaesthetic visit were satisfied, compared to 62.5% of those who did not have a preoperative visit, with this association being statistically significant (p-value = 0.001). Participants with no prior history of using anaesthesia had a higher satisfaction rate (81.8%) compared to those with a prior history of anaesthesia use (67.6%), and this association was also statistically significant (p-value = 0.001). Furthermore, it was found that patients classified as ASA three had lower satisfaction (35.7%) compared to those of ASA one (85.8%) and ASA two (75.1%), with the association being statistically significant. Additionally, the provision of explanations about anaesthesia was significantly associated with satisfaction with perioperative anaesthesia care, with the patients who received explanations about anaesthesia being more satisfied (67.5%), compared to those who did not receive (15.8%) explanation about anaesthesia-Table 2.

**Table 2:**
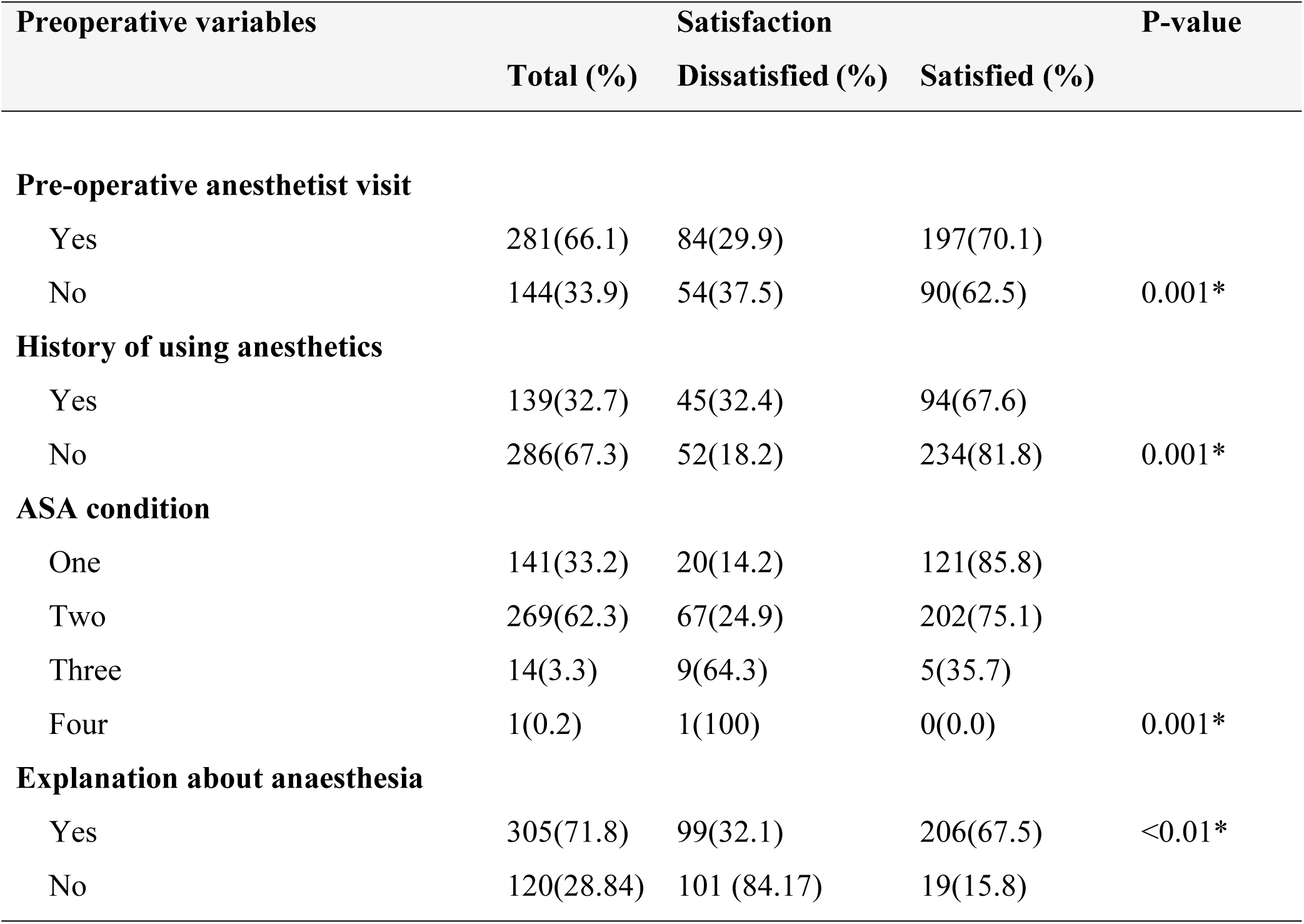
Distribution of preoperative factors across patients’ satisfaction with perioperative anesthesia care among general surgical patients (N = 425).

Among study participants, only 66.1% were visited by the anesthetist in the preoperative period. 71.8% reported that the anesthetists had provided explanations about anaesthesia. Only 32.7 % of study patients had a history of previous anesthesia exposure-figure 2.

**Figure 1:**
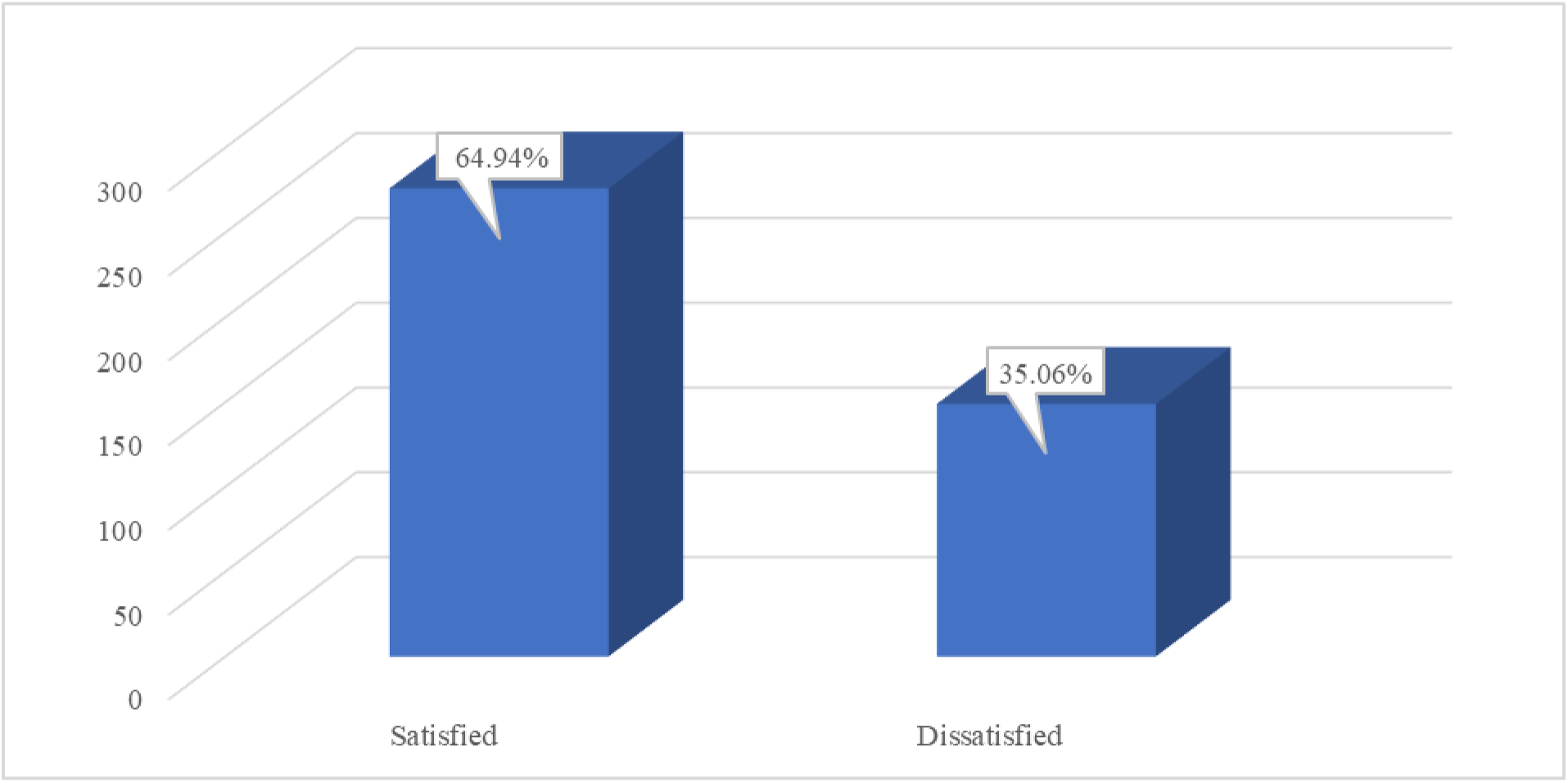
Overall patient satisfaction with perioperative anesthesia care among surgical patients admitted at Dodoma Regional Referral Hospital, Tanzania.

**Figure 2:**
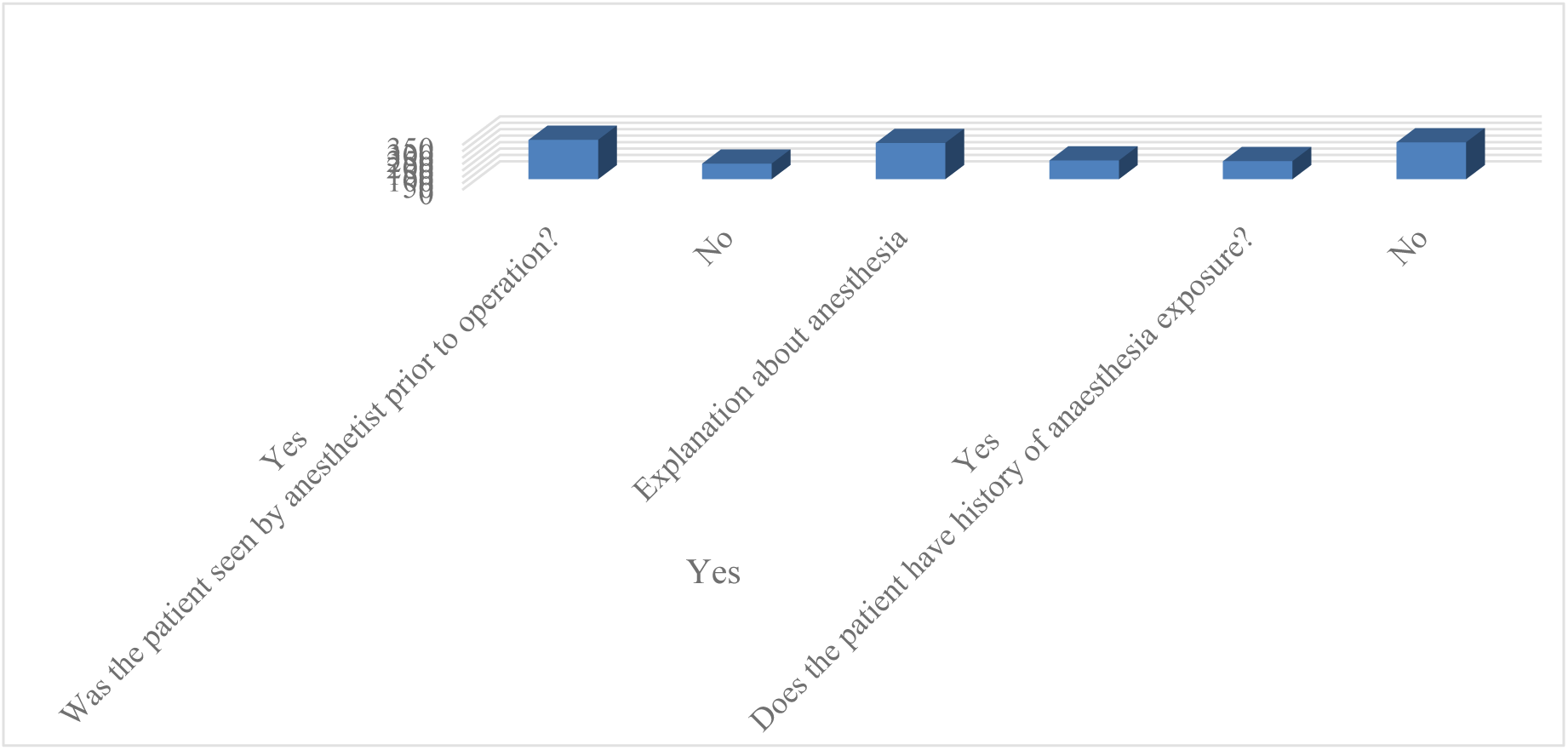
Shows the Preoperative factors of general surgical patients who have undergone surgery at Dodoma Regional Referral Hospital (N=425).

Intraoperative factors were also distributed across patients’ satisfaction with perioperative anesthesia care. Table 3 shows that emergency surgeries predominated, accounting for 69% of cases, and the majority of patients (81.4%) underwent major surgeries. However, there was no statistically significant association between the type of surgery and patient satisfaction. Additionally, 69.3% of patients who received premedication reported being satisfied with the anaesthesia care, compared to only 59.5% of those who did not receive premedication, with this association being statistically significant. There was no significant association between the type of anaesthesia administered and patient satisfaction (see Table 3).

**Table 3:**
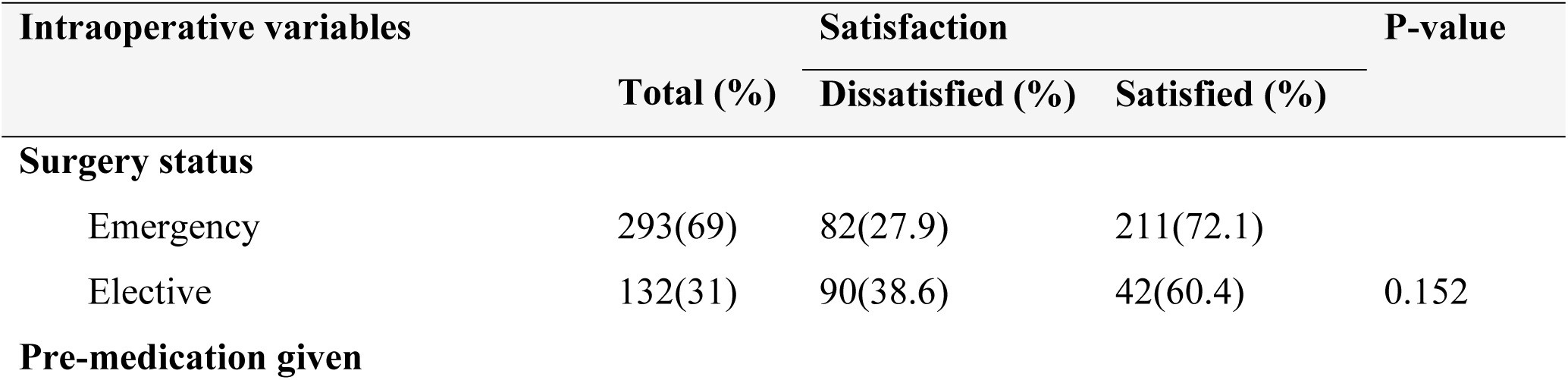
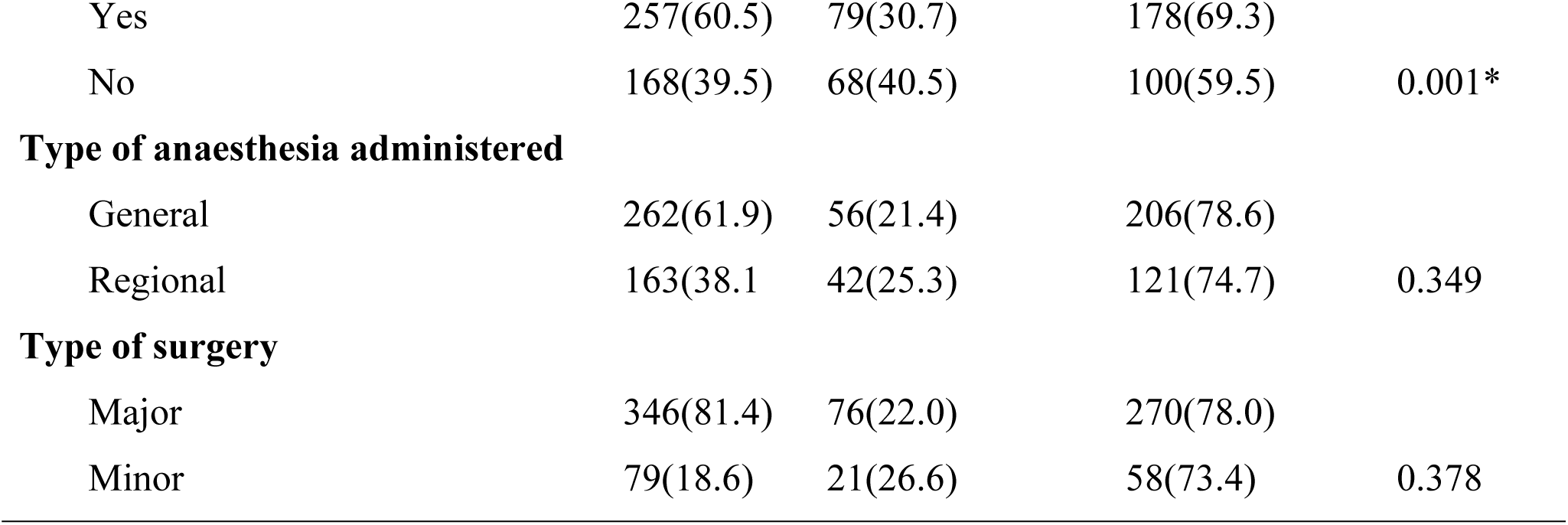
Distribution of Intraoperative factors across patients’ satisfaction with perioperative anesthesia care among general surgical patients (N = 425).

Table 4 shows the distribution of intraoperative factors across satisfaction among general surgical patients who have undergone surgery under regional anaesthesia at Dodoma Referral Regional Hospital. For patients who received regional anesthesia during intraoperative time, there was a significant association between pain experienced during surgery and overall satisfaction. Similarly, privacy status was significantly associated with patient satisfaction; the majority of patients whose privacy was respected (56.9%) were satisfied, compared to those whose privacy was not respected (46.15%). There was a statistically significant association between privacy status and satisfaction

**Table 4:**
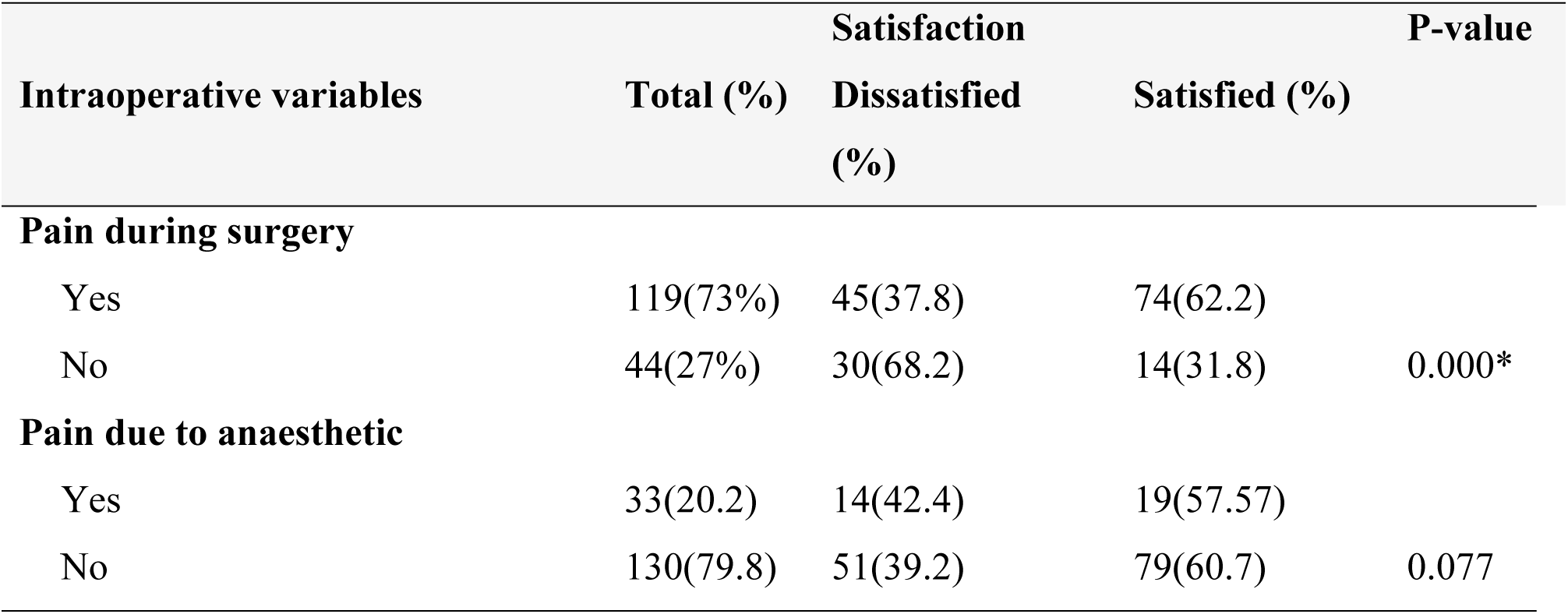
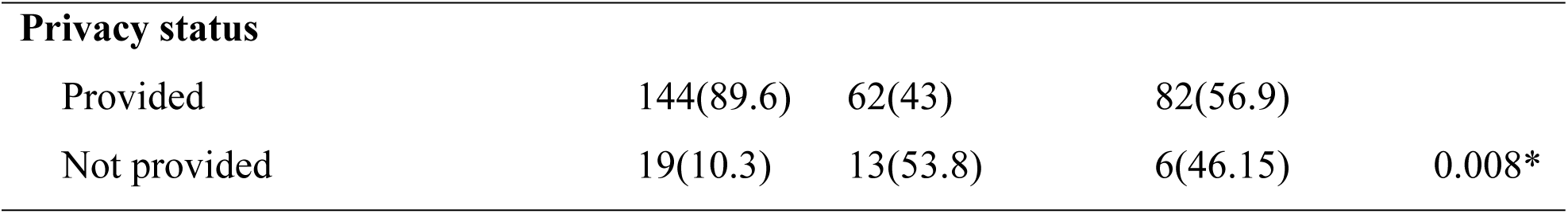
Distribution of intraoperative factors across satisfaction among general surgical patients who have undergone surgery under regional anaesthesia at Dodoma Regional Referral Hospital, (N = 163).

To determine the group difference, the study distributed postoperative factors across patients’ satisfaction with perioperative anesthesia care. We found that among the study respondents, 64% of those who experienced postoperative surgical site pain and 72.9% of those who did not experience postoperative surgical site pain were satisfied with their anaesthesia care, with surgical site pain being significantly associated with satisfaction. Additionally, the majority of patients (56.8%) who spent a shorter time in the recovery room were satisfied, compared to 28.8% of those who spent a longer time, with this association being statistically significant.

For patients who underwent surgery under regional anaesthesia, there was a statistically significant association between postoperative headache and satisfaction, with those experiencing headaches being less satisfied (37.5%) compared to those who did not experience headaches (58.1%)-table 5 below.

**Table 5:**
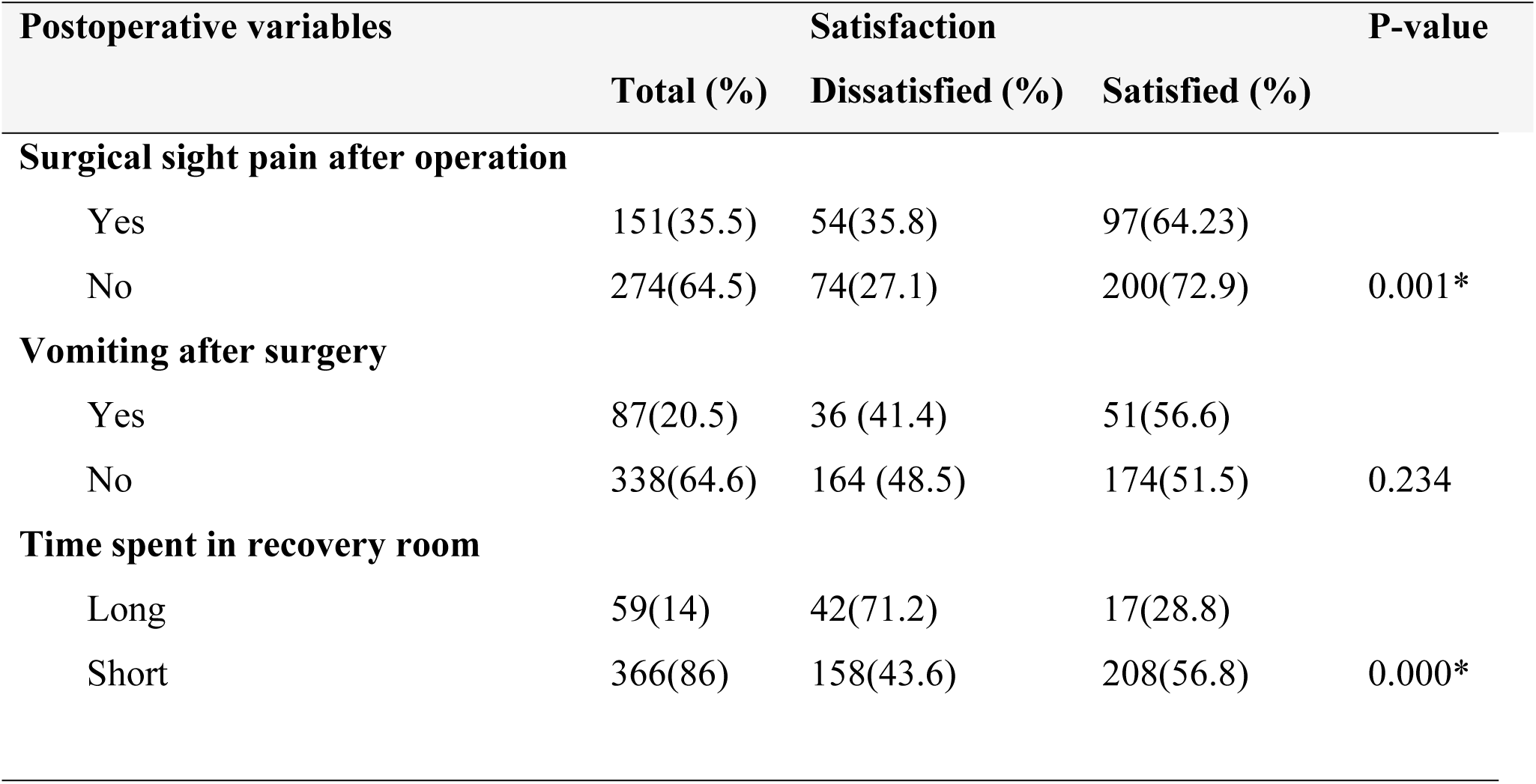
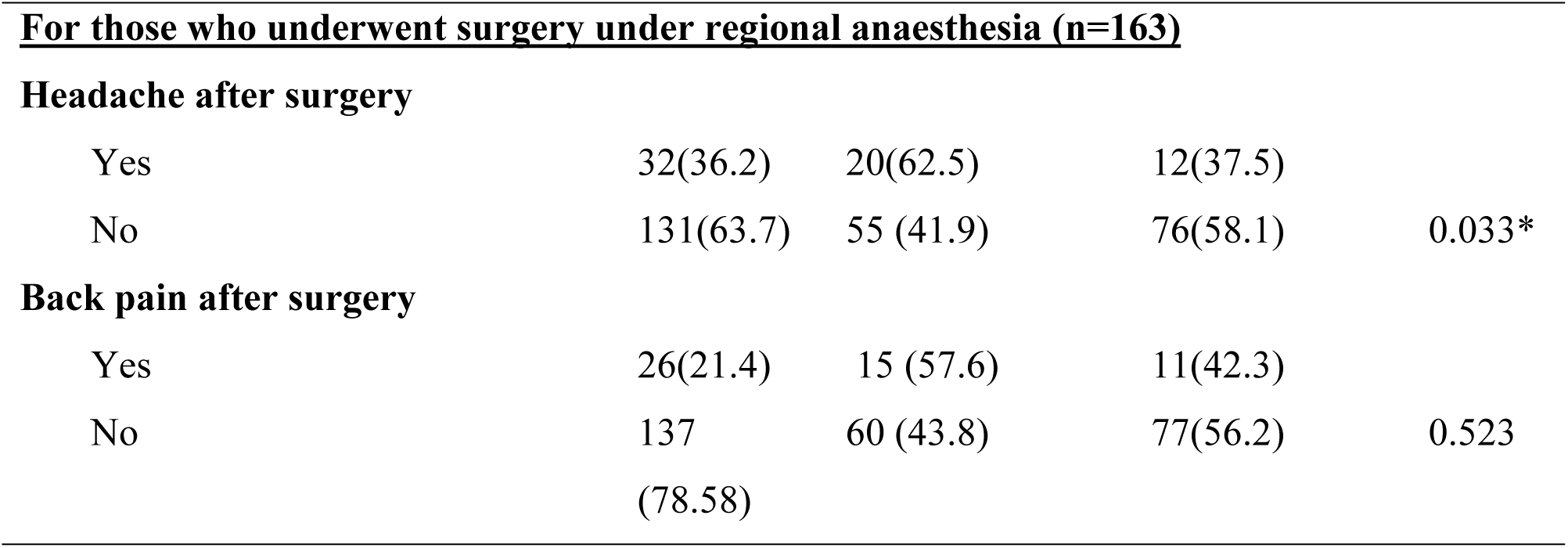
Distribution of postoperative factors across patients’ satisfaction with perioperative anesthesia care among general surgical patients (N = 425).

The majority, 73%, were willing to return to hospital for other operation services, 3.5 % refused to return to hospital and 23.5 % were not sure if they could return or not. Figure 3: below illustrates the information.

**Figure 3:**
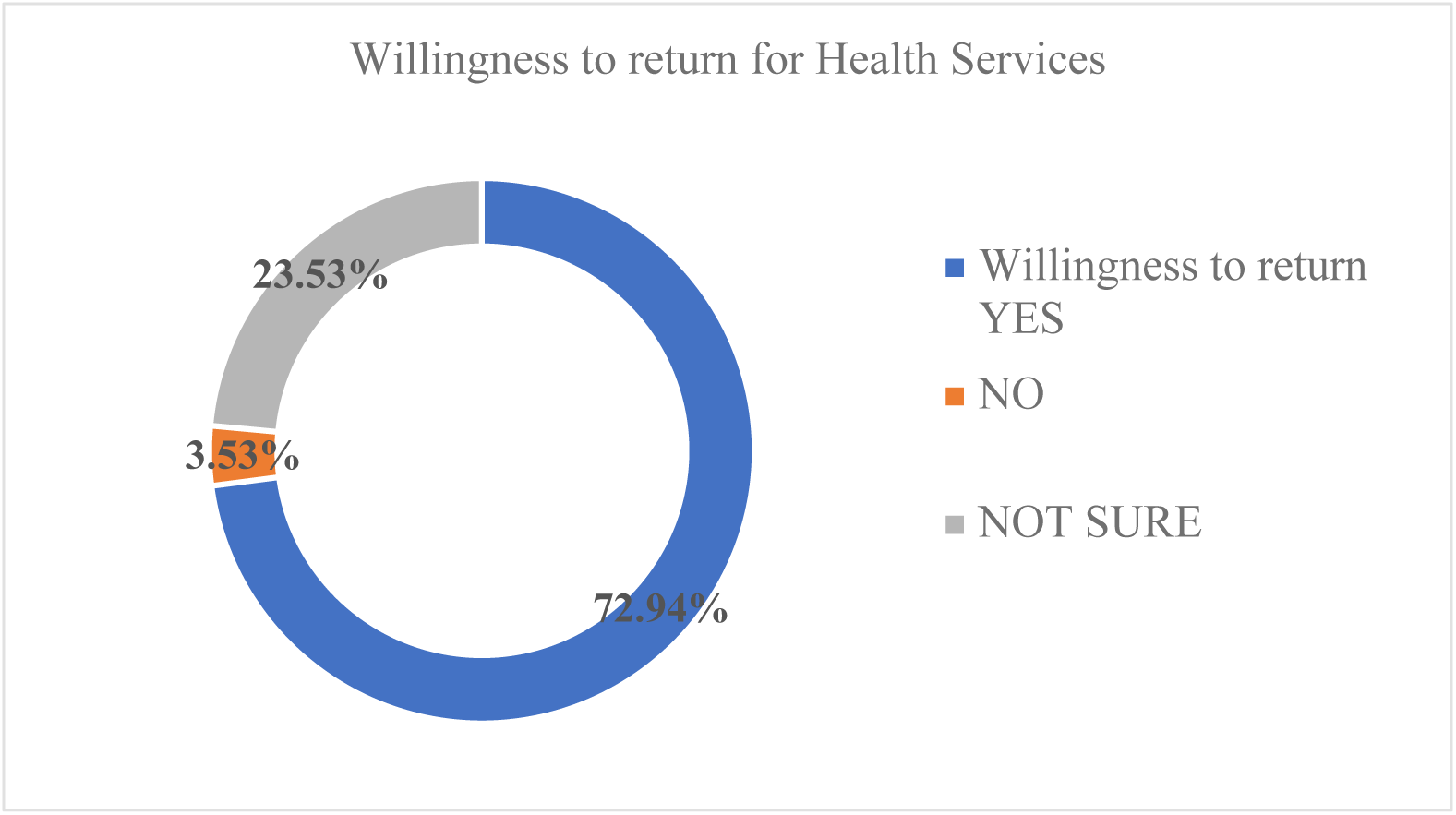
The proportion of patients who are willing to return to the same hospital following the perioperative anaesthesia.

Table 6 presents the multivariate logistic regression results. The results shows that patients with whom the preoperative visit was conducted were more likely to be satisfied (AOR 1.9, 95% CI 1.09-3.6, P-value 0.023) compared to those whose preoperative visit was not conducted. In terms of ASA classification, respondents with ASA three were less likely to be satisfied (AOR 0.05, 95% CI 0.006-0.44, P-value 0.007) compared to those with ASA (four) class. Patients in ASA two class were more than one time likely to be satisfied (AOR 1.9,95% CI 0.31-0.99, P-value 0.02) compared to patients in ASA four.

**Table 6:**
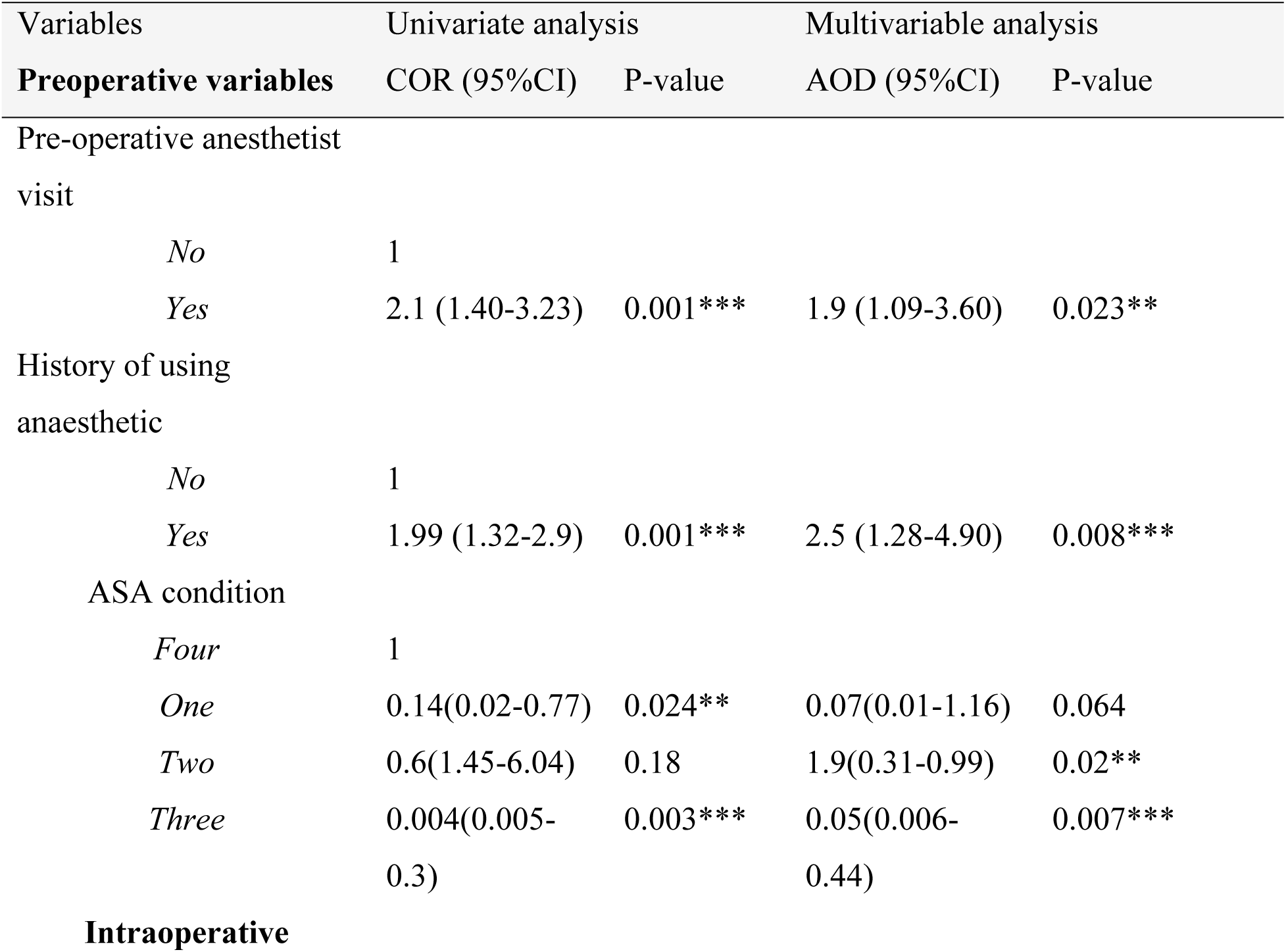
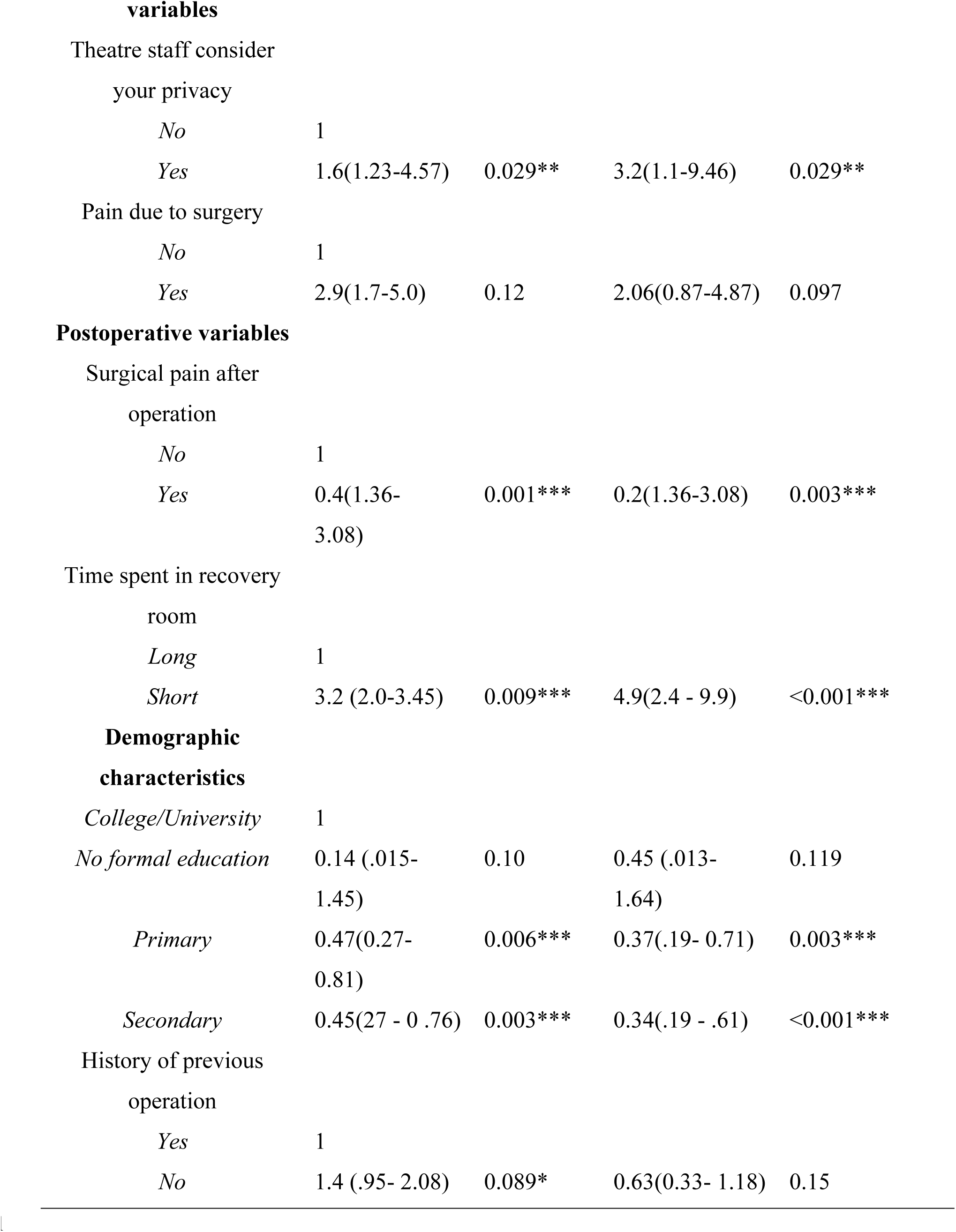
Multivariable logistic regression of factors associated with satisfaction of participants (N = 425).

Regarding theatre staff considering patients’ privacy, patients whose privacy was considered were more than three times likely to be satisfied (AOR 3.2, 95%CI 1.1 – 9.46, P-value 0.007) compared to those whose privacy was not considered. Those who experienced post-operative pain were 80% less likely to be satisfied (AOR 0.2, 95% CI 1.36-3.08, P-value 0.029), compared to those who did not experience postoperative pain.

In terms of time spent in the recovery room, those who spent less time were more than four times likely to be satisfied (AOR 4.9, 95% CI 2.4 - 9.9, P-value <0.001) compared to those who spent longer time in the recovery room. The results above have taken into consideration the control variables (i.e level of education and history of previous operations).

## DISCUSSION

The present hospital-based study involved general surgical patients who underwent anaesthesia care and were admitted at Dodoma Regional Referral Hospital aimed at determining patients’ satisfaction with perioperative anaesthesia care and its associated factors among these patients.

In the present study the overall patient satisfaction score was relatively low compared to the one calculated from the demarcation threshold formula .However this finding was closely similar to a study conducted in Eretria and Saudi Arabia[5,15]. Possible explanation might be similarities in socio-demographic profiles, such as age distribution, education levels, and economic status, which can influence patient expectations and perceptions of healthcare services. Further, utilization of the same assessment tools and similar healthcare infrastructure, including the availability of medical equipment, facilities, and resources dedicated to perioperative care. In contrast to this study, a study conducted in UK and Australia found a higher satisfaction of patients in the perioperative period.[16,17]. This difference could be due to more advanced infrastructure, advanced pain management protocols and techniques and emphasis on patient-centered care, which focuses on meeting individual patient needs, preferences, and values.

The relatively low overall patient satisfaction score with perioperative anesthesia care found in this study suggests that there are significant areas for improvement in the perioperative care provided. Factors contributing to this lower satisfaction score likely include issues in preoperative preparation, intraoperative management, and postoperative care. It is crucial for the hospital to address these areas comprehensively to enhance the patient’s experience and outcomes. A cross-sectional study reported that gender is an independent risk factor for patient satisfaction, with males being more satisfied, which aligns with the findings of this study [4,6]. However, in the present study, gender was not a significant associator of patient’s satisfaction. A possible reason for this discrepancy could be the difference in analytical methods. Previous studies primarily used descriptive analysis, while in this study, we conducted logistic regression analysis to test the strength of the association between the dependent and independent variables.

This study found that patients being pre-visited by anesthetists before operation, history of anaesthesia exposure and ASA status would significantly influence patients’ satisfaction. Preoperative visits by anesthetists significantly increased patient satisfaction by providing vital information, addressing concerns, and building rapport, suggesting that routine preoperative visits should be implemented. Patients with previous anesthetic exposure reported higher satisfaction due to reduced anxiety and a better understanding of the process, highlighting the need for thorough patient education and preoperative counselling, especially for first-time surgical patients. Findings were similar to those reported by Makoko et al from South Africa[18].

Additionally, the ASA status of patients significantly impacted satisfaction, with healthier patients being more satisfied, indicating the importance of tailored perioperative care plans for patients with higher ASA statuses to improve their experiences and outcomes. This is contrary to the results reported by Fetene et al which showed ASA was not significantly associated with satisfaction[4], indicating a need for a tailored approach to patients’ severity which is the case in Eritrea[5]. A study conducted by Sun reported that patients with a history of anesthesia exposure were more satisfied than those without prior exposure[1], findings that align to these study findings.

The study found that pre-medication provision was significantly associated with satisfaction. Studies have reported that preventing postoperative nausea and vomiting is a significant factor for perioperative patient satisfaction following anaesthesia care, [4,19,20] which aligns with the present study. In our study, premedication provision was significantly associated with satisfaction. .Additionally, a study in Saudi Arabia found that emergency procedures had greater odds of patient satisfaction than in elective surgeries [8,15].

Findings from this study indicated that pain due to surgery was a major contributor to patient satisfaction for those who underwent regional anesthesia, a result not commonly observed in other similar studies from Ethiopia and Saudi Arabia[4,8]. This highlights the importance of considering methodologies for adequate intraoperative pain management.

In this study, experiencing postoperative pain was found to be significantly associated with patient dissatisfaction, consistent with the findings of previous research[21]. Postoperative pain is a critical determinant of overall patient satisfaction with perioperative anesthesia care. Effective pain management not only enhances patient comfort but also reduces the risk of chronic pain development and accelerates recovery.

Studies have consistently shown that inadequate pain control can lead to negative postoperative experiences, resulting in decreased patient satisfaction[1,18]. Therefore, it is important to prioritize pain management strategies tailored to individual patient needs to improve overall satisfaction with anaesthesia care.

Furthermore, a significant factor influencing patient satisfaction in this study was the time spent in the recovery room. Prolonged recovery room stays can be distressing for patients, potentially leading to increased anxiety and discomfort. Efficient management of the recovery process is essential to ensure a smooth transition from anesthesia to full recovery. Previous studies have indicated that lengthy recovery room times are often associated with increased pain, nausea, and overall discomfort, which negatively impact patient satisfaction. By optimizing recovery room protocols and ensuring prompt and effective postoperative care, healthcare providers can enhance patient experiences and satisfaction levels[17].

This study found that majority of patients are willing to return for future procedures following their perioperative anesthesia care. Many studies emphasize that negative perioperative experiences can lead patients to share their bad experiences with friends and relatives, discouraging them from returning to the facility [1,15,22] none have specifically examined the willingness to return after anesthesia care. The findings form a baseline for other studies in the improvement of the perioperative care. This finding is particularly significant as the willingness to return serves as a critical indicator of overall patient satisfaction and confidence in the healthcare services provided.

The study has identified significant areas for improvement in preoperative preparation, intraoperative management, and postoperative care. Notably, factors such as preoperative visits by anesthetists, prior anesthesia exposure, and ASA status were key influences on satisfaction, highlighting the need for improved patient education and communication in the perioperative setting.

### Study strength, limitation and mitigation

This study has a significant strength in that it provided a thorough evaluation of patient satisfaction, covering multiple aspects of the perioperative anaesthesia experience, from preoperative information to postoperative recovery, and provides valuable baseline data that can be used for future research and comparisons, helping to track improvements and changes over time.

Further, this study assessed the willingness to return, which reflects the level of patient satisfaction, an aspect not previously explored in similar studies.

One limitation of this study was collecting data before participants were discharged which was the potential restraint on them from expressing their thoughts due to their dependency on care. This challenge was addressed by conducting an exit interview. Additionally, we provided assurance, built a strong rapport throughout the study, engaged in clear communication, demonstrated empathy, and proactively addressed their concerns.

Conducting the study at one Centre (i.e Dodoma Regional Referral Hospital) restricts the generalizability of the findings to other hospitals with different healthcare settings and patient demographics. To address this limitation, we increased the sample size to include a broader population, enhancing the study’s generalizability.

## Conclusion

The overall perioperative satisfaction with anesthesia care at Dodoma Regional Referral Hospital is relatively low compared to the Leiden Perioperative Care Patient Satisfaction and some previous studies. However, higher proportion of patients expressed the willingness to return for similar care at the hospital, indicating a baseline of patient satisfaction that warrants further investigation to understand the reasons behind this acceptance.

Key factors significantly associated with perioperative satisfaction include respect for patient privacy during surgery and effective postoperative pain management. Additionally, preoperative anesthesia visits and patients’ ASA status also influence satisfaction levels. To enhance patient satisfaction, factors contributing to dissatisfaction must be addressed, and active involvement from all stakeholders is crucial.

Future research should consider including patients’ willingness to return as a key measure of perioperative satisfaction and investigate the reasons behind patient dissatisfaction that includes evaluation of other aspects of patient satisfaction and discharge criteria.

## Ethical consideration

Ethical clearance for the study was obtained from the MUHAS Research and Publication Committee (IRB reference number DA.282/298/01.C/2226), and permission to conduct the research was granted by Dodoma Regional Referral Hospital. The study prioritized the human rights of respondents by ensuring their autonomy, with informed consent sought from all participants. Patients who chose not to participate still received full treatment, and those experiencing distress during the interview were given care before continuing. Confidentiality was maintained by ensuring that no patient identities were disclosed in the final report. Sensitive data was handled with care to protect participants’ privacy.

## Data availability

Data are kept at the MUHAS data repository site, and they are available upon reasonable request.

## Acknowledgment

The authors would like to thank the Dodoma Regional Referral Hospital management, data collectors, and study participants for their contributions to this study.

## Author Contribution

JM- Designed the study, wrote methodology, data curation, data analysis, wrote original draft of manuscripts, funding acquisition.

AA- Conceptualization of the study, methodology, reviewing of manuscripts drafts, overseeing the entire research process

AK- Conceptualization of the study, methodology, reviewing of manuscripts drafts, overseeing the entire research process

AU- Reviewed the first manuscript and added updates to the manuscript

FS-Contributed to the data collection, analysis and reviewing the initial draft of the manuscript

IN – Investigation and data curation

*All authors have read and agreed to the published version of the manuscript.

